# Incremental Learning Approach for Semantic Segmentation of Skin Histology Images

**DOI:** 10.1101/2025.10.07.25336648

**Authors:** Sana Fatima, Anum Abdul Salam, Muhammad Usman Akram, Ibrahim A. Hameed, Saad B. Ahmed

## Abstract

This study presents an incremental learning framework to enhance the generalization and robustness of transformer-based deep learning models for segmenting skin cancer and related tissue structures. While deep learning models often perform well on data distributions similar to their training sets, their accuracy typically degrades when exposed to novel scenarios—limiting their clinical utility in skin cancer diagnosis. To address this, we propose a biologically inspired incremental learning strategy tailored for skin cancer classification and segmentation, allowing the model to incorporate new data progressively while reducing catastrophic forgetting. Our approach integrates multiple loss functions to preserve existing knowledge while adapting to additional magnification levels. Experimental results on the in-distribution test set demonstrate consistent performance improvements: achieving 89.05% accuracy with 10× magnification, 92.68% with 10× and 5× combined, and 95.53% when incorporating 10×, 5×, and 2× magnifications. These findings highlight the potential of our method to improve the adaptability and reliability of deep learning systems for empirical generalization in skin cancer classification tasks.

**Please note:** Abbreviations should be introduced at the first mention in the main text – no abbreviations lists. Suggested structure of main text (not enforced) is provided below.

## I. Introduction

Evaluating the advances and shortcomings related to the Sustainable Development Goals (SDGs) today is vital for decision makers seeking to improve population health [1]. One of the latest estimates is that the global burden of cancer will continue to increase during the next two decades [2]. Skin cancer is one of the most common diseases on the globe. There are two general skin cancer types: non-melanoma skin cancer and melanoma skin cancer. In the United States, non-melanoma skin cancers (NMSCs), such as squamous cell carcinoma (SCC) and basal cell carcinoma (BCC), make up over 98% of all skin cancer cases [3]. The Cancer Society of America states that melanoma skin cancer makes up just 1% of all cancer cases, and its death rate is higher [4]. A timely and precise diagnosis is still essential for successful treatment [5].

### A. Skin Cancer Diagnosis

To correctly identify and treat skin cancer, pathologists examine several tissue slides that have different histomorphological traits. For differentiation among different histological layouts, all excision specimens must be focused, cut into 3 mm sections, and dyed with hematoxylin and eosin (H&E) [6], [7]. However, earlier approaches that digitize skin cancer diagnosis using binary classification provide limited clinical and economic value, as they offer less informative outputs compared to multi-class semantic segmentation. In contrast, segmentation-based methods can deliver more detailed insights, enhancing decision-making confidence for both professionals and patients, where accurate detection and diagnosis are critical. While multi-class semantic segmentation methods offer more detailed reports suitable for clinical decision-making, binary classification methods often lack this level of clarity. Furthermore, Fig 1 shows the regions that are the most affected by skin cancer according to global statistics, with North America representing approximately fifty percent of all instances [8].

**Fig. 1.**
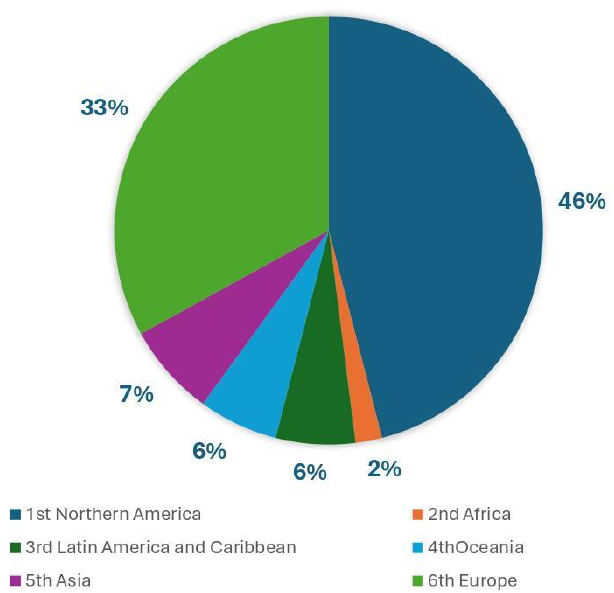
Global distribution of skin cancer rate.

Our proposed research uses histology images, and microscopic images of tissues used to analyze the disease, particularly in cancer diagnosis. Using the histology images, we proposed an incremental learning semantic segmentation of a multi-class framework on NMSC. Our study’s contribution is as follows;

- We propose an innovative incremental learning algorithm that uses Segformer as the baseline model with an emphasis on the semantic segmentation of NMSC and tissues.
- This strategy combines various loss functions, incorporating novel learning with knowledge distillation (KD) to avoid catastrophic forgetting by transfer of knowledge from a previously trained model (teacher) to a new model (student), along with joint training for both novel and previously known classes.
- Furthermore, a mutual distillation loss function is included in an incremental semantic segmentation approach, which leads to improved performance in real-world applications.

## II. Literature Review

The widespread acceptance of artificial intelligence (AI) in clinical diagnostics, particularly for skin cancer, depends not only on algorithmic performance, but also on the development of reliable and adaptive models [9]. Although various machine learning (ML) and deep learning (DL) techniques have shown great accuracy in detecting skin cancer, the majority of them depend on static training data and are not well suited to adjust to new types of lesions or updated datasets [8], [10].

Traditional models based on DL do not adequately represent the requirement for continual learning [11]–[13], which has led to the development of incremental learning (IL) as explained in [14]. Once trained, these models require extensive retraining to integrate new classes of leison, which is computationally costly and prone to catastrophic forgetting [15], [16].

A significant problem with IL is the occurrence of catastrophic forgetting, in which previously learned information is wiped out. Solutions like Knowledge distillation (KD) and memory-aware loss functions are effective techniques [17], [18]. Wu et al. provided an integrated loss strategy for incremental classification that retained performance without utilizing previous data. However, most IL techniques have focused on classification, and few have investigated their incorporation into semantic segmentation, particularly in healthcare.

Recent research focuses on incremental approaches to semantic segmentation. Michieli et al. [19] proposed the IL problem solving for semantic segmentation, focusing on pixel-level labeling. Cermelli et al. [20] proposed weakly incremental learning for semantic segmentation (WILSS), which employs weak supervision to introduce additional segmentation classes. These approaches demonstrate the possibility of IL for segmentation, but they have not been extensively tested on histopathological images or multiclass skin cancer segmentation, which is the primary objective of our research. Recent advances have used transformer-based models, which are recognized for successfully capturing global context in vision tasks. Xie et al. [21] developed Segformer, a transformer-based segmentation model that is lightweight and highly efficient. Although Segformer has been used in a variety of medical fields, its potential for incremental multiclass skin cancer segmentation has yet to be explored. Our study addresses this gap by transforming Segformer into an IL method for non-melanoma skin cancer (NMSC) segmentation, using KD and mutual distillation methods to reduce forgetting.

Other research has concentrated on employing lightweight encoders or improved CNNs to increase segmentation accuracy. Kosgiker et al. [22] proposed SEGCAP (a capsule network architecture for object segmentation), which improves the precision of the lesion boundary in noisy pictures. While these approaches produce good static efficiency, they cannot adapt in real-time to novel lesion forms that IL provides.

Furthermore, Imran et al. [23] presented a transformer-based approach to identify NMSC using histopathological images within an incremental learning architecture. While their approach indicates the importance of transformers in IL instances, it is primarily concerned with classification tasks and does not dive into semantic segmentation or multiclass lesion distinction, both of which are essential for clinical diagnosis and treatment planning. However, our proposed method applies incremental learning to semantic segmentation, using the Segformer architecture and incorporating a unique mutual distillation loss. This enables our model to maintain existing information despite learning new classifications of lesions and improves spatial localization, which is vital in clinical processes.

Multiclass segmentation remains challenging [24] due to similarities between classes and data imbalance. Moradi et al. [25] proposed a multiclass visual segmentation technique for skin cancer images that uses a hybrid dictionary approach, illustrating adequate performance on a limited dataset. However, such techniques often require additional feature design or retraining. Our technique addresses these difficulties by using a single transformer-based IL pipeline that allows incremental class addition without the need for manual intervention.

In conclusion, while previous research has improved segmentation and IL independently, few studies have combined these areas in the context of multiclass skin cancer detection using histopathology images. Our contribution is to provide an IL-based semantic segmentation mechanism using Segformer that can learn new skin cancer classifications over time while retaining existing information. This significant development focuses on clinical demands for flexible diagnostic algorithms that can evolve in tandem with new data without having to retrain from scratch.

## III. Methodology

In this section, we discuss the methodology we used to analyze histological images. The following section details the proposed architecture.

### A. Segformer Architecture

Our segmentation approach is powerful, fast, and robust, allowing for greater automation while getting computationally efficient [21]. We used segformer [21], our baseline model. The transformer-based architecture has two modules: encoder and decoder, as shown in Fig 2.

**Fig. 2.**
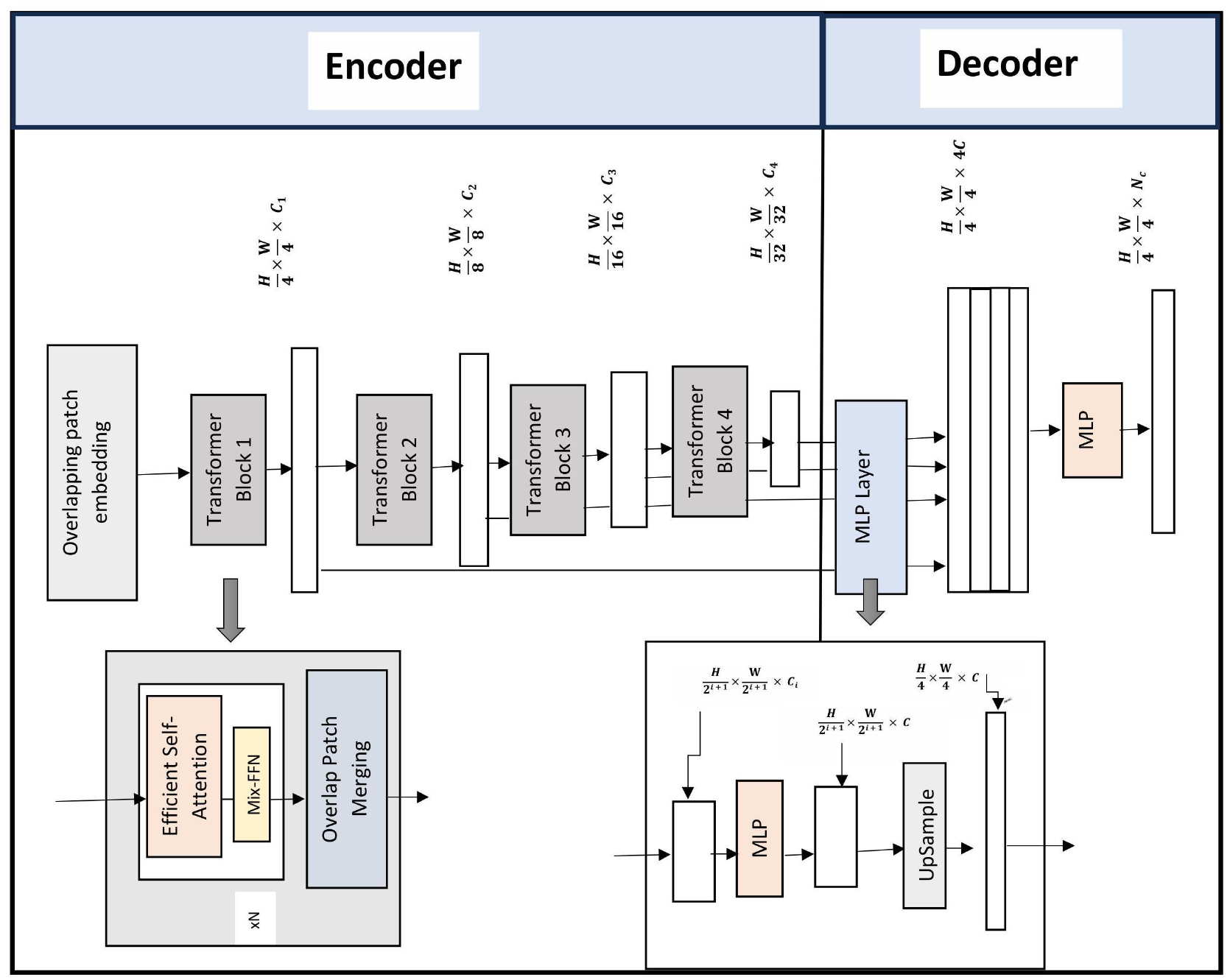
The Segformer architecture is used as a backbone in our proposed framework. The Segformer design consists of an encoder and a decoder. The encoder comprises overlapping patch embedding, transformer blocks, and efficient self-attention methods. The decoder combines MLP and upsampling layers to generate better semantic segmentation outcomes. While the fundamental Segformer concept is used as is, our innovations are in combining this backbone with an incremental learning approach based on mutual distillation (described in Section 3.2).

The encoder’s self-attention mechanism (SAT) is essential as it enables the model to identify long-distance connections between various image regions. SegFormer improves segmentation performance by effectively collecting both local and global information through using self-attention as depicted in Fig 3.

**Fig. 3.**
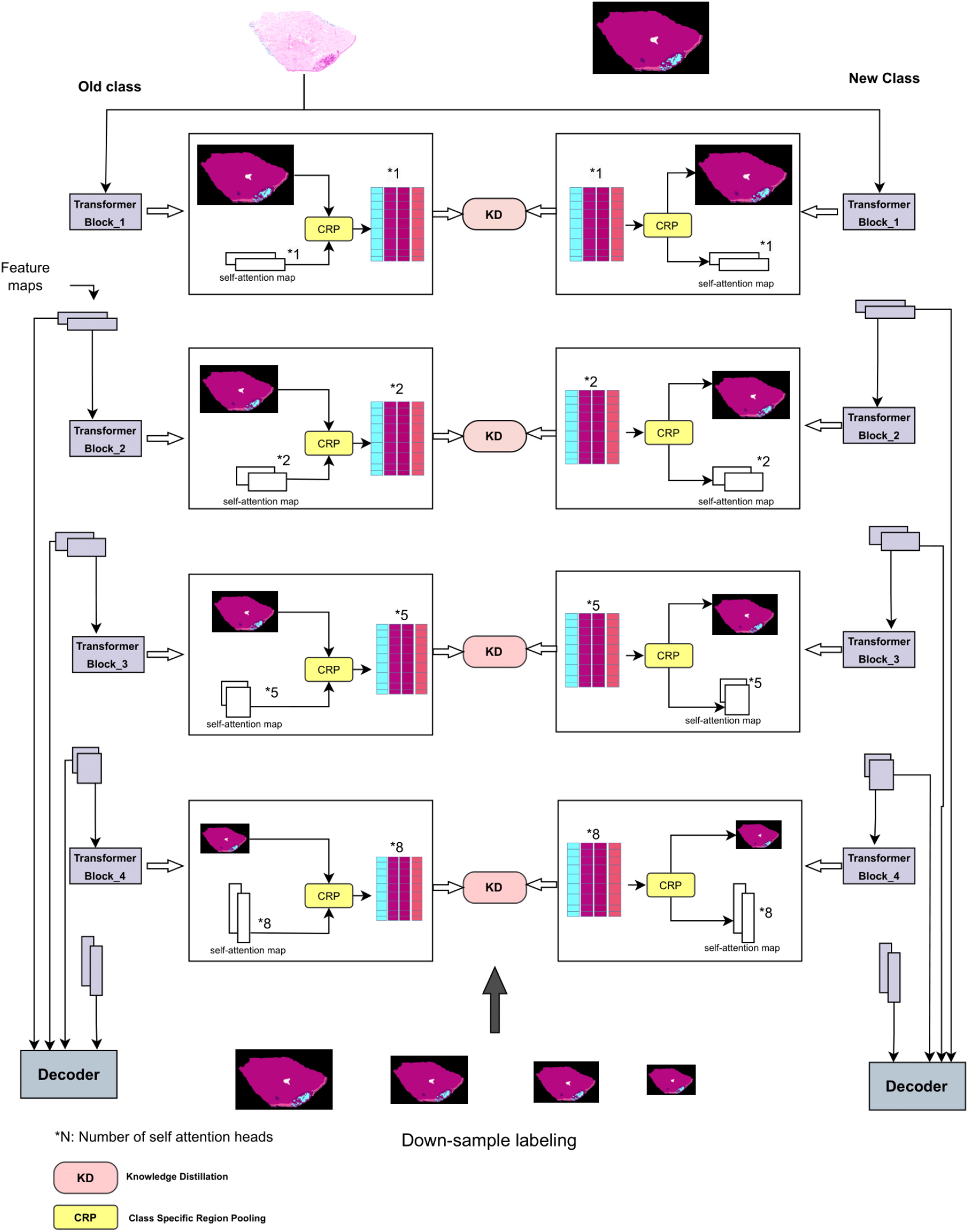
Design of a self-attention transfer (SAT) approach for incremental semantic segmentation. In every transformer encoder block, self-attention maps have been extracted from the final multi-head self-attention layer. During incremental segmentation, these self-attention matrices are utilized for knowledge distillation, enabling learning within and between classes.

#### 1) Encoder

The encoder is an organized hierarchical structure to acquire coarse and fine attributes with both high and low resolution. The decoder module uses a multi-layer perceptron (MLP) that incorporates features from the multilayer encoder to offer semantic segmentation of the input image.

The method generates 4 × 4 tiles from an input image with H × W × 3, where H and W refer to height and width. The size of the 4 × 4 patch is appropriate for tasks requiring segmentation with dense features. The 4 × 4 patches are integrated into a hierarchical encoder that acquires four different features depending on the input image size. The MLP decoder utilizes the multi-level extracted features to produce a segmentation of size 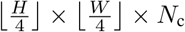, where *N*_c_ denotes the total number of subcategories that need to be segmented.

#### 2) Architecture of Proposed Mix Transformer Encoders

We adopt a sequence of Mix Transformer encoders (MiT) having the same architecture and varied sizes, from MiT_B0 to MiT_B5. MiT_B0 is a light model for rapid prediction, while MiT_B5 is an extended model for optimal performance. MiT design depends on a vision transformer (ViT) [27] but is improved for semantic segmentation. MiT_B0 to MiT_B5 features include the depth, the number of heads, embedding measurements, and several parameters are represented in TableI.

**TABLE I.** A Characteristics of Mix Transformer Encoder Models.

| Model | Number of Depth (Layers) | Embedding Dimension | Quantity of Heads | Parameters (Millions) |
| --- | --- | --- | --- | --- |
| MiT-B0 | 4 | [32, 64, 160, 256] | [1, 2, 5, 8] | 3.4 |
| MiT-B1 | 4 | [64, 128, 320, 512] | [1, 2, 5, 8] | 13.1 |
| MiT-B2 | 4 | [64, 128, 320, 512] | [1, 2, 5, 8] | 24.2 |
| MiT-B3 | 4 | [64, 128, 320, 512] | [1, 2, 5, 8] | 45.6 |
| MiT-B4 | 4 | [64, 128, 320, 512] | [1, 2, 5, 8] | 60.2 |
| MiT-B5 | 4 | [64, 128, 320, 512] | [1, 2, 5, 8] | 81.2 |

Unlike the ViT model, which produces a single-resolution features map, this module is meant to generate multi-level features using the input image, which are comparable to those generated by CNNs. Such multiple-level characteristics preserve either high-resolution images coarse interpretations or low-resolution fine-grained knowledge, which are vital to enhancing semantic segmentation tasks. Patch merging is used to generate the hierarchical feature map *F*_*i*_with a resolution of 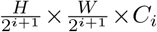, where *i* ∈ {1, 2, 3, 4} and *C* and is greater than *C*_*i*_. This hierarchical technique allows for the analysis of multi-scale characteristics across various resolutions.

The patch merging method applied to ViT merges [21] a *N* × *N* × 3 patch with an image patch to create a 1×1 ×*C* vector. A hierarchical mapping of features can be generated by extending this technique to combine a 2 × 2 × *C*_*i*_ feature patch into a 1× 1 × *C*_*i*+1_ vectors. In addition, we can apply an iterative approach to each feature map within the hierarchy, reducing our hierarchical characteristics to *F*_1_ (with dimensions 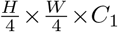) to *F*_2_ (with dimensions 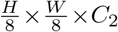).

Nonetheless, the original goal of this method was to integrate non-overlapping sections of an image or feature, which could lead to an elimination of local coherence surrounding these patches. To overcome this limitation, we utilize an overlapping patch merge technique. We indicate three parameters: *M, N*, and *O*. The patch size is expressed as *M*, the stride between consecutive patches is indicated by *N*, and the padding size is indicated by *O*. To achieve overlapping patch merging while retaining feature sizes comparable to those achieved by the non-overlapping approach, we chose *M* = 7, *N* = 4, and *O* = 3 as one configuration in the tests we performed, and *M* = 3, *N* = 2, and *O* = 1 for other in our configuration.

### a) Efficient Self Attention

Our SRA, similar to multihead attention (MHA), gets a query q, a key k, and the value v as input and produces a refined feature. The following is an expression for the SRA:

The equation for SRA(q, k, v) is

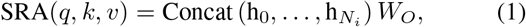

where

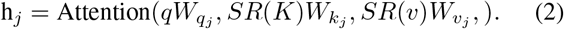

Concat(·) represents the concatenation operation of multiattention heads. The coefficients 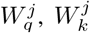, and 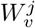 are proportional projection with dimension 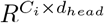, and *W*_*O*_ has dimension 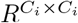. *N*_*i*_ represents the number of heads in the attention layer of Stage *i*, while the dimensions of each head, *d*_*head*_, is determined by *C*_*i*_*/N*_*i*_. Although it decreases the spatial dimensions of *k* and *v* before the attention computation, the Spatial Reduction Attention (SRA) [26] operates similarly to the MHA.

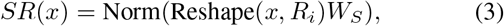

Here, 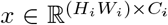 represents the initial sequence, and *R*_*i*_ is the reduction ratio for Stage *i*. The *Reshape*(x, *R*_*i*_) operation alters the dimensions of *x* to 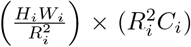. The coefficient matrix 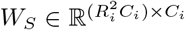 transforms the input sequence to *C*_*i*_ dimensions. The **Norm(·)** function refers to layer normalization.

The self-attention layer in the encoder structure is the primary cause of the computational bottleneck. Here is the way the self-attention function is calculated:

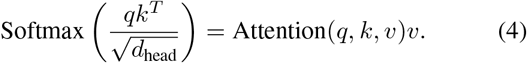

where, the query vectors (q), key vectors (k) and value vectors (v) in the traditional multi-head self-attention mechanism all have identical dimensions: *N* × *C*, where *N* = *H* × *W* indicates the sequence length.

The computational complexity of this approach using equations (1, 3) is *O*(*N* ^2^), making it unfeasible at high image resolutions. We apply the order reduction method explained in [28] to address this issue. The sequence length is reduced using a reduction ratio *r* in this process, resulting in the following transformations: The expression.

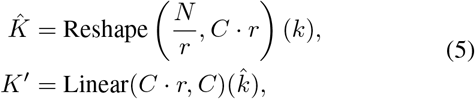

where *k* is the sequence to be decreased, Reshape 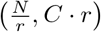 (*k*) reshapes *k* to have dimensions 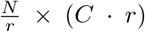, and Linear(*C*_in_, *C*_out_)(*·*) indicates a linear transformation that transforms a *C*_in_-dimensional tensor to a *C*_out_-dimensional tensor. Consequently, the dimensions of the new *K* are 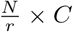. With this modification, the self-attention mechanism’s complexity is effectively reduced from *O*(*N* ^2^) to 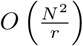.

The effective length of the sequence decreases as *r* grows, which lowers the computational cost and immediately impacts the sequence length reduction. We set *r* in our tests to be to [64, 16, 4, 1] for each of the stages 1 through 4.

### b) Mix-FFN

Transformer-based models often use positioning encoding (PE) to acquire spatial data about input attributes. However, if the dimensions of training and test images vary, this technique may result in a performance decrease as well. In such cases, the fixed-resolution Positional Embedding (PE) [29] has to be transformed, which frequently results in reduced model performance. To overcome this challenge, conditional positional encoding (CPE) [30] provides a driven-by-data positional encoding generator (PEG) that can deal with durations longer than those for training models. To implement PEG using a two-dimensional convolution with a 3 × 3 kernels when used with PE. This method relies on the premise that convolutional neural networks (CNNs) intrinsically encode positional information utilizing zero padding and boundaries. Segformer claims that explicit PE is insufficient for semantic segmentation problems.

Segformer utilizes a mixed feedforward network with a 3 x 3 convolution kernel, as shown as follows:

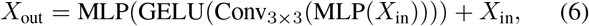

The efficient self-attention module’s extracted features are expressed by *X*_in_. Combining MLP with 3 × 3 convolution in Mix-FFN provides output enhanced with spatial information, shown in equation (6).

#### 3) Decoder Architecture

SegFormer offers a relatively small decoder based entirely on MLP layers, that reduces the need for the complex and highly computational elements employed by previous methods. This decoder becomes simplified by its hierarchical Transformer encoder, which offers a larger effective receptive field (ERF) than conventional CNN-based encoders.

The recommended all-MLP decoder includes four primary stages. To establish uniform channel dimensions, the MiT encoder multilevel features (*F*_*i*_) are transmitted through an MLP layer. The features are subsequently upsampled to one-fourth of their original dimension and combined. The third stage involves adding encapsulated features (F) to another MLP layer. A multilevel processing (MLP) layer subsequently processes the fused feature to predict the segmented mask *M* with a resolution of 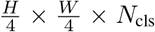, whereby *N*_c_ shows the number of classes. The full procedure of the decoder may be described as follows:

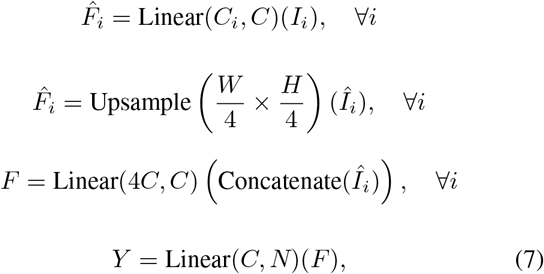

Here, Y is the predicted segmented mask, and Linear(*C*_in_, *C*_out_)( ·) indicates a linear layer with input and output dimension *C*_in_ and *C*_out_, accordingly.

### B. Learning without forgetting

To overcome catastrophic forgetting while allowing for multiscale learning, the proposed approach uses incremental learning for semantic segmentation. This method enables the model to adapt to novel data distributions while keeping previously learned knowledge a critical capacity for whole slide image (WSI) segmentation at multiple resolutions. As new resolutions are introduced, relevant self-attention qualities from prior training phases are retained using knowledge distillation as shown in Fig 3.

Unlike other techniques such as domain adaptation, self-supervised learning, and meta-learning, we selected the incremental route because it allows magnifications to be incorporated stepwise rather than subjecting extensive retraining over previous data. Unlike domain adaptation, which requires simultaneous access to both source and target domains (difficult for high-resolution WSIs), or self-supervised and meta-learning methods, which are either task-misaligned or computationally intensive, incremental learning allows for the stepwise integration of magnifications without retraining on prior data. This combines flexibility, generalization, and effectiveness, making it ideal for medical image segmentation applications.

Data incremental learning enables the model to progressively adapt to new information while maintaining performance on previously seen data, improving its generalization across varying data distributions. Fig 4 illustrates the data incremental learning technique. Initially, the algorithm is trained on data with a certain resolution, it learns from both new and prior resolutions, by keeping focus on new ones. This method allows the model to evolve data requirements over time. Importantly, the evolution to data incremental learning is adaptable, according to the introduction of new classes or changes in data specification.

**Fig. 4.**
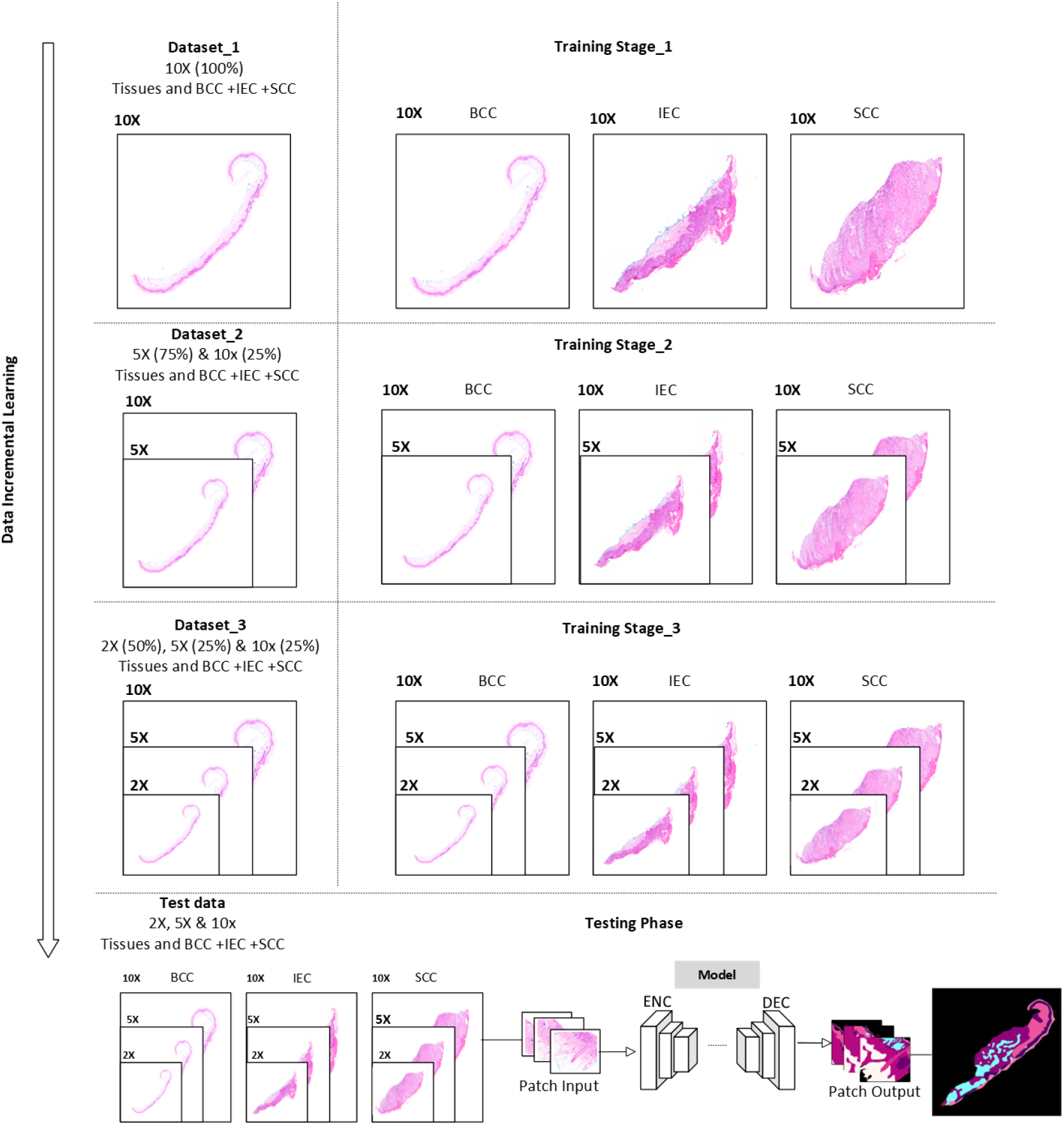
The training steps illustrate the data incremental learning strategy along the axis that is vertical. The testing stage of a model displays the evaluation set, which comprises all resolution-based data.

We gradually train our model, gaining newly acquired information while keeping the previously obtained information. To address the difficulty of catastrophic forgetting, we employ a hybrid strategy that involves mutual distillation loss. This method offers efficient knowledge distillation by replaying previous data throughout each incremental training session. At each stage, the training data contains the newly offered samples from the same classes, ensuring balanced retention and continual learning while adjusting to evolving data distributions. As seen in Table II, the training results were obtained by incorporating new and already trained data. The technique uses a process with three stages in which data of various resolutions (10x, 5x, and 2x) are used differently at each level. In Stage-1, uses 100% of the data at 10x resolution. Stage-2 provides data a 5x resolution, which accounts for 75% of the total, whereas only 25% of 10x data is maintained. Finally, in Stage-3, use 50% of the data at a resolution of 2x, as well as 25% at a resolution of 10x and 5x.

**TABLE II.** Proposed Framework Incremental Training Strategy and Respective Stages.

| Data Resolution | Stage – 1 | Stage – 2 | Stage – 3 |
| --- | --- | --- | --- |
| 10x | 100% | 25% | 25% |
| 5x | - | 75% | 25% |
| 2x | - | - | 50% |

Class imbalance, especially in rare diseased tissues, is a prevalent problem in skin histology data. Our incremental learning technique addresses this by introducing the model to samples at various magnifications, reducing the influence of underrepresented classes. Furthermore, rotation-based augmentation (90°, 180°, 270°) was given to rare classes, increasing their presence. This strategy balances training data and improves segmentation efficiency in rare tissue types.

#### 1) Knowledge distillation and new learning

Knowledge distillation is a model reduction and acceleration approach that employs a teacher model’s guidance to improve the accuracy of smaller models [31]. This technique helps improve the accuracy of the student model while maintaining computational efficiency. In the context of incremental learning, knowledge distillation plays a crucial role in mitigating catastrophic forgetting by preserving essential information from previous learning stages while allowing the model to adapt to new data. During the training stage, we use a combination of two loss functions: classification loss *L*_cls_ and mutual distillation loss *L*_mdl_ to address specific challenges. The loss of incremental semantic segmentation *L*_incremental_ is stated as:

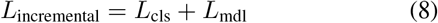

### a) Classification Loss

The classification loss *L*_cls_ is determined using categorized cross-entropy loss, which is frequently utilized in the semantic segmentation roles. The classification loss equation,

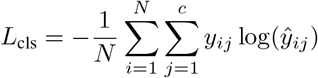

The classification loss is expressed as *L*_cls_, where *N* denotes the total quantity of instances and *c* the total amount of categories. The actual label of instances *i* and class *j* is *y*_*ij*_, while *ŷ*_*ij*_ is the projected likelihood that sample *i* relates to class *j*. The factor 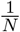 ensures that the loss is averaged in every sample.

As our dataset incorporates old and new samples from the same classes, the algorithm integrates its prior knowledge of categories while acquiring novel ones. The following equation represents the incremental classification loss:

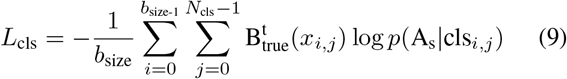

where,

- *b*_size_: shows the batch size.
- *N*_cls_: reflects the total number of classes acquired for the current task.
- 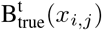: includes labels from recently acquired classes.
- *x*_*i,j*_: the input training sample in the current task (*i, j*).
- A_s_: relates to the representations of already learned classes.
- *p*(A_s_ | cls_*i,j*_): represents the model’s predicted probability for the true class.
- cls: shows the classes in the dataset.

The input for every phase of incremental learning includes color images, designated as *x* ∈ 𝕏^2^, which comprise the training data for the same classes with parameter specifications.

### b) Mutual Distillation Loss

The mutual distillation loss *L*_mdl_ employs Bayesian inference to evaluate structural and semantic similarities among the original and incrementally acquired representation [31]. By optimizing these distributions together, the model ensures that while gaining new knowledge, it maintains previously learned information. The loss is determined by corresponding the variances of the previous and new representations to retain information during incremental learning as follows in equations (10, 11).

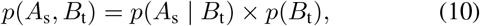

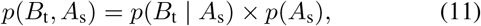

As equation (10) represents the relationship between old (*A*_s_ student) and new (*B*_t_ teacher) representations with conditional probability. The equation (11) represents the inverse relationship between the old (student) ones impacting the new (teacher) representations. B_t_ and A_s_ indicate the feature representations of newly acquired and previously acquired knowledge, accordingly.

By including such distributions in the model training process, the model can understand the connection between new and original knowledge. This method guarantees that the model acquires new information properly while maintaining previously retained knowledge.

This loss evaluates the disparity between the model’s prediction likelihood distribution and the actual label distribution. By decreasing the KL divergence, the predictions made by the model become more closely related to the true labels. The equations (10, 11), represent the KL divergence of these two probabilities, which could be related to the mutual distillation loss: The KL divergence between *p*(*A*_s_) and *p*(*B*_t_) is as follows:

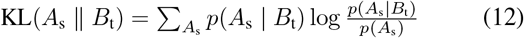

Also, the reverse KL divergence among *p*(*B*_t_) and *p*(*A*_s_), are:

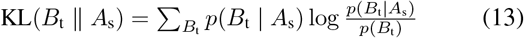

Subsequently, the total of these two KL divergences is the loss of mutual distillation:

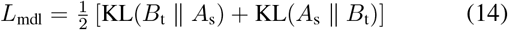

### c) Final Objective Function

To efficiently train our proposed incremental learning system, we integrate the classification loss and mutual distillation loss into a final objective. The final loss function *L*_total_ is defined as:

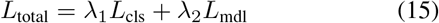

The hyperparameters *λ*_1_ and *λ*_2_ balance each loss component’s contribution. These hyperparameters were experimentally selected based on performance on the validation set to guarantee consistent learning and knowledge retention. This ensures that the test set remained unseen during tuning, preserving the integrity of the evaluation. The final aim solves the catastrophic forgetting problem that comes with incremental learning by making sure the model learns to categorize new classes and extract information from previously seen data at the same time.

#### 2) Joint Training

To improve the learning of acquired data through joint training, we propose that training data at every incremental step include instances from both old and new sub-data. Since the model has previously learned the old data and maintains prior information via knowledge distillation, the data for the new resolution will make up the majority of the training set. Notably, our technique is not a memory constraint. However, we recommend utilizing a limited quantity of unseen data from previous data for re-learning.

## IV. Experimental Implementation

### A. Dataset

The dataset underlying this study is composed of 290 histopathology slides of Queensland University dataset [32], each one of which has been annotated by a pathologist to determine the tissue sections that are most usual of non-melanoma skin cancer. The dataset consists of BCC 140 slides, SCC 60 slides, intraepidermal carcinoma (IEC) 90 slides, containing biopsy specimens collected via shave (100), punch (58), and excision (132) process. These slides, examined throughout four months from late 2017 to early 2018, contain patients ranging in age from 34 to 96, an average age of 70, and a ratio of men to women is 2:1. Utilizing a camera (DP27 Olympus microscope) at 10x the magnification, overlapped tiles were merged into high-resolution images mosaics with a pixel size of 0.67 *µ*m. Images ranged from 11 million to 500 million pixels.

### B. Ground Truth Segmentation

Specimen slides indicated twelve classification categories, including carcinomas. These include the following: Squamous Cell Carcinoma (SCC), Background (BKG), Basal Cell Carcinoma (BCC), Reticular Dermis (RET), Glands (GLD), Hair Follicles (FOL), Inflammation (INF), Hypodermis (HYP), Intraepidermal Carcinoma (IEC), Papillary Dermis (PAP), Keratin (KER), and Epidermis (EPI). The dataset was annotated and categorized by dermatologists, and pathologists collaborated to generate pixel-level annotations, which took approximately 250 hours. Furthermore, they categorize the dataset into 12 classes, represented by pixel color information as shown in Fig 5.

**Fig. 5.**
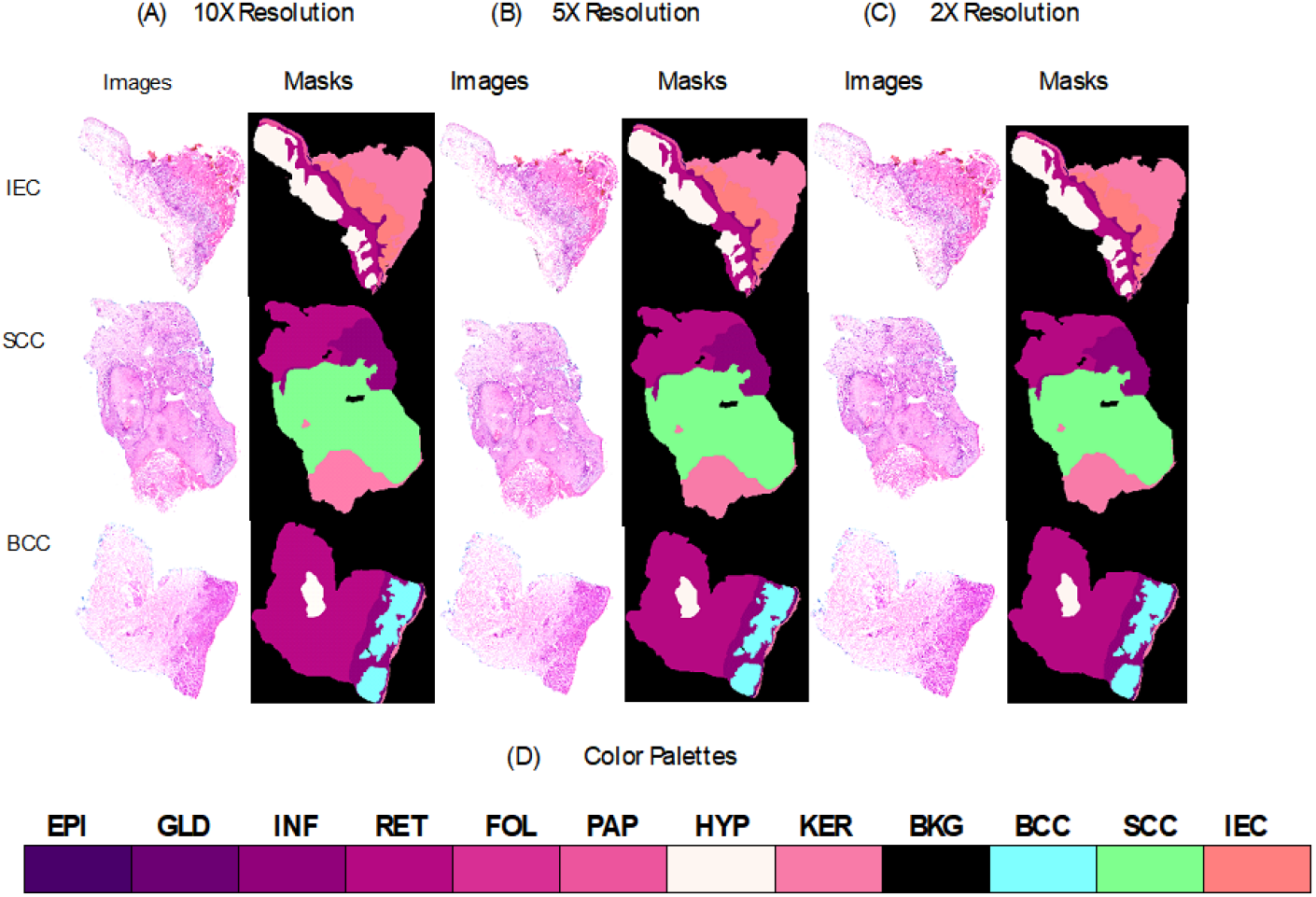
Non-melanoma skin cancer (NMSC) such as SCC, IEC, and BCC at different resolutions with corresponding ground truth masks and color palettes. (A) Specimens at 10X resolution with masks. (B) Specimens at 5X resolution with masks. (C) Specimens at 2X resolution with masks. (D) Multi-class color palette.

The diversity in certain classes, such as the epidermis, which shows a high degree of non-cancerous variation, presented a significant challenge for creating these segmentations. To address this, the ideal healthy epidermis has been referred to as EPI, while slices with dysplastic keratinocytes and carcinomas were categorized as IEC. Certain segmentation boundaries, such as those among the EPI and PAP dermis, were visible at high magnification, whereas others, such as those between the PAP and RET dermis, were less clear. In addition, pixel-wise classification accuracy provoked uncertainty regarding the maximum achievable performance, as 100% ground truth correctness cannot be ensured [23]. To compare, the dataset was downsampled by factors such as 2x, 5x, and 10x, showing that the 2x dataset maintained sufficient tissue attributes for analysis, but greater downsampling (e.g., 10x) transformed the interpretability of tissue borders as depicted in Fig 5.

### C. Training Specifics

The Segformer as a base model used in the proposed work is implemented in Python 3.11.4. To train the model, We used incremental segmentation loss as specified in equation (8). The overall loss *L*_incremental_ is built by integrating the classification loss (*L*_cls_), and the mutual distillation loss (*L*_mdl_) respectively.

The baseline model was trained using the facility provided by the Digital Research Alliance of Canada, in addition to the NVIDIA MIT_B4 model. However, for the incremental learning framework, we use 8GB NVIDIA GeForce RTX 2070. The model was trained using 50 epochs with a batch size of 2. It used the Adam optimizer with a learning rate of 0.0006. The normalizing layers 1 × 10^*−*6^ were employed for better generalization, particularly for lower batches. The original images were far too large to fit straight into the network, consequently overlapping patches were generated. We split the data into three sets i.e., testing, validation, and training, with a ratio of 10:10:80. Each patch was sized at 256 × 256 pixels. We also applied color the color jitter augmentation during training. For incremental learning, we used dataset resolutions of 2x, 5x, and 10x, as well as testing and validation at the same resolution. To fix class imbalances throughout training, patches containing these categories were enhanced using rotation in 3 angles (90,180,270), improving their presence fourfold.

The dataset was divided using stratified sampling to ensure that all tissue types were represented proportionally in the training, validation, and test sets at every magnification level (10x, 5x, and 2x). The data was separated into three categories: 80% for training, 10% for validation, and 10% for testing. Stratification helps to reduce class imbalance, resulting in a fair and uniform model evaluation.

## V. Results and Discussion

### A. Initial Results

In our studies, we used the Segformer Mit_B4 framework as the baseline for incremental learning semantic segmentation. The characteristics of this framework are shown in Table I. We trained the Segformer model on three different resolution-bases,ets that are 10x, 5x, and 2x. This resulted in three distinct trained models, each verified against resolution-specific validation data. Then, all models were tested at all resolutions. Table III illustrates the performance evaluation of suggested framework.

**TABLE III.** Performance Comparison between Baseline Model and Proposed Model on Test Dataset Multiple Resoluions.

| Data Resolution | Model | Accuracy | IoU | DSC |
| --- | --- | --- | --- | --- |
| 10x | SegFormer (Baseline) | 89.5% | 80.2% | 88.9% |
| 5x |  | 84.1% | 74.5% | 85.4% |
| 2x |  | 80.9% | 70.6% | 82.8% |
| 10x, 5x | Proposed Framework | 92.68% | 85.3% | 92.1% |
| 10x, 5x & 2x |  | **95.53%** | **88.7%** | **94.0%** |

From Table III, the highest performance was achieved by the Segformer model trained on the 10x data resolution, having an average accuracy of 89.5%, an IoU of 80.2%, and a DSC of 88.9%. The 5x resolution model has 84.1% accuracy, 74.5% IoU, and 85.4% DSC. The 2x resolution model has the lowest, with 80.9% accuracy, 70.6% IoU, and 82.8% DSC. These findings suggest that higher-resolution images have greater spatial detail, leading to improved segmentation ability. In contrast, the suggested incremental learning framework (trained successively on 10x, 5x, and 2x) produced the highest overall results, with 95.53% accuracy, 88.7% IoU, and 94.0% DSC, showing its effectiveness in robust multiresolution segmentation.

### B. Ablation Studies

Using Segformer as a baseline, we developed the incremental learning method shown in Fig 4. We used an incremental learning method to train all model layers sequentially across phases. The results obtained from this framework are summarized in Table III.

The gained results indicate that our proposed methodology is effective across several phases of incremental learning when applied to histopathology data. In Stage-1, a pre-trained Segformer model used 10x resolution data and achieved an accuracy of 89.50%. Subsequently, in Stage-2, the data incremental learning technique, which included new (5X) and previously viewed data, increased accuracy to 92.67%. Using knowledge distillation, the model efficiently preserved existing information while adding new input. In Stage-3, adding 2x resolution data to lower amounts of 10x and 5x resolution data improved the model’s performance, resulting in an accuracy of 95.53%. These findings show that by combining incremental learning with knowledge distillation, the model may retain past information while adjusting to new input, resulting in higher performance.

To further show the viability of our strategy, we provide the visual results of the proposed framework in Fig 7, which corresponds to Table III. The figure shows improved prediction accuracy, especially for test samples with 5x and 2x resolutions. These visualizations highlight the ability of the model to generalize effectively across resolutions while preserving past information, which is essential for the successful segmentation of histopathology data.

To assess the efficiency of the proposed framework, we examined class distribution and accuracy in a variety of tissues, such as BCC, IEC, SCC, and other tissues. The violin plot Fig 6 compares the class distributions of the baseline and suggested frameworks, demonstrating improved balance and representation.

**Fig. 6.**
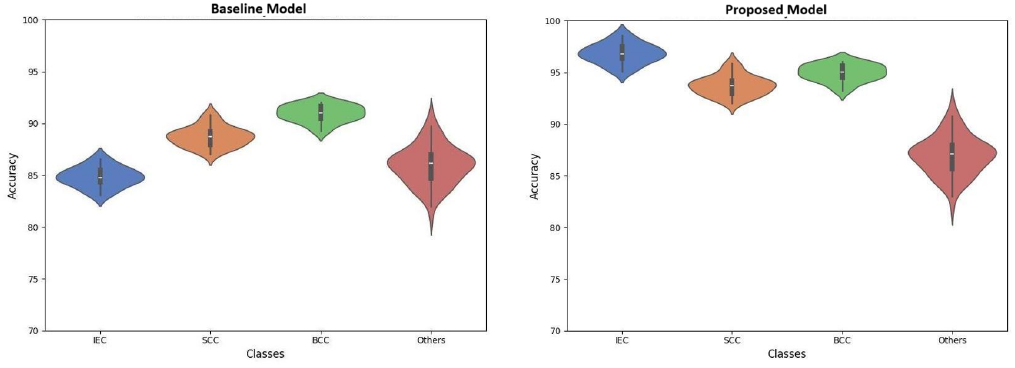
The violin plot shows the class distribution across BCC, IEC, SCC, and other tissues for both the baseline and the suggested framework, emphasizing the improved class balance produced by the incremental learning strategy.

**Fig. 7.**
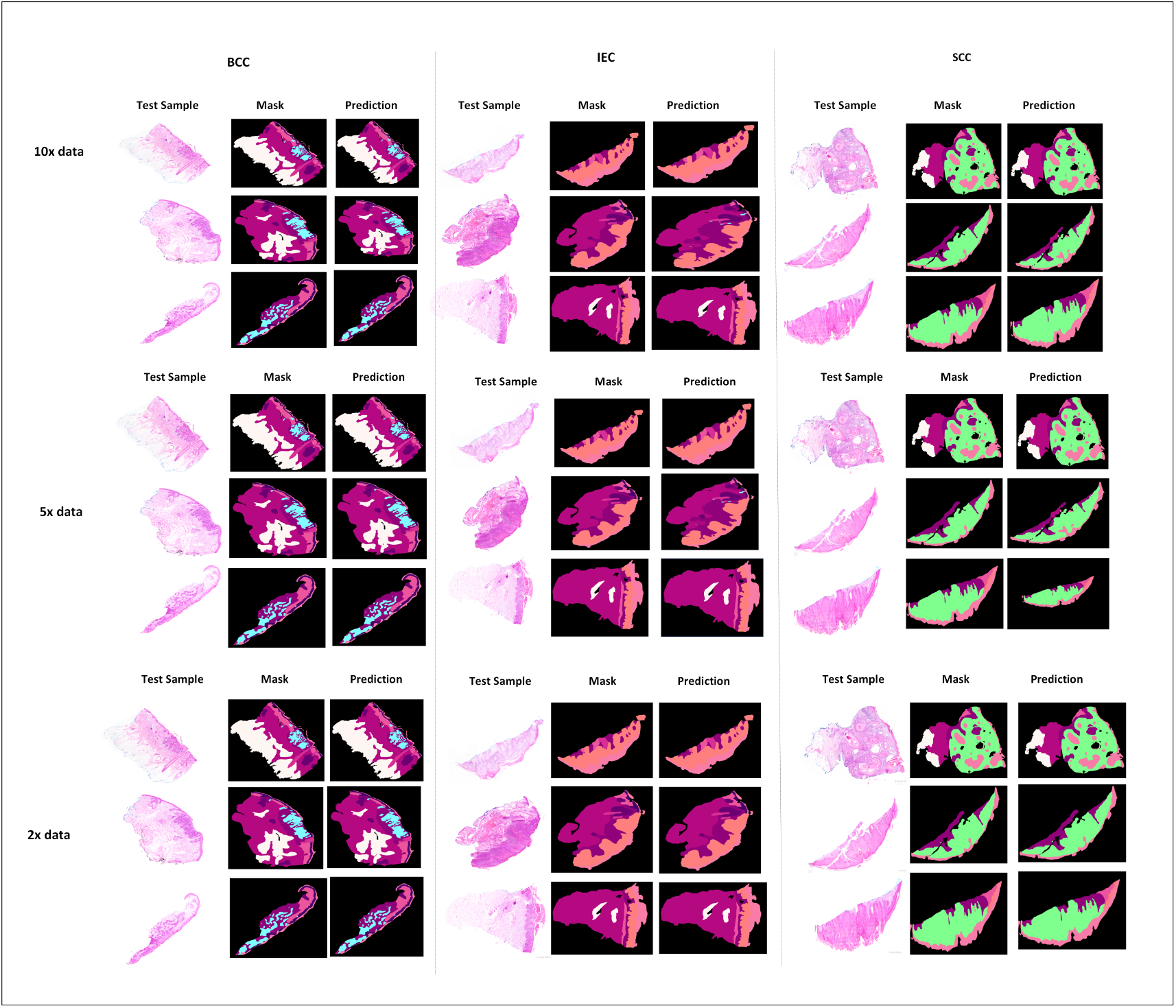
Multi-resolution images of non-melanoma skin malignancies such as IEC, BCC, and SCC are shown together with ground truth and predictions produced using the data incremental learning framework.

The proposed approach improves prediction performance for BCC, IEC, SCC, and mixed carcinoma cases, as seen in the confusion matrices in Fig 8. These findings show the model’s adaptability and generalizability across a variety of test settings.

**Fig. 8.**
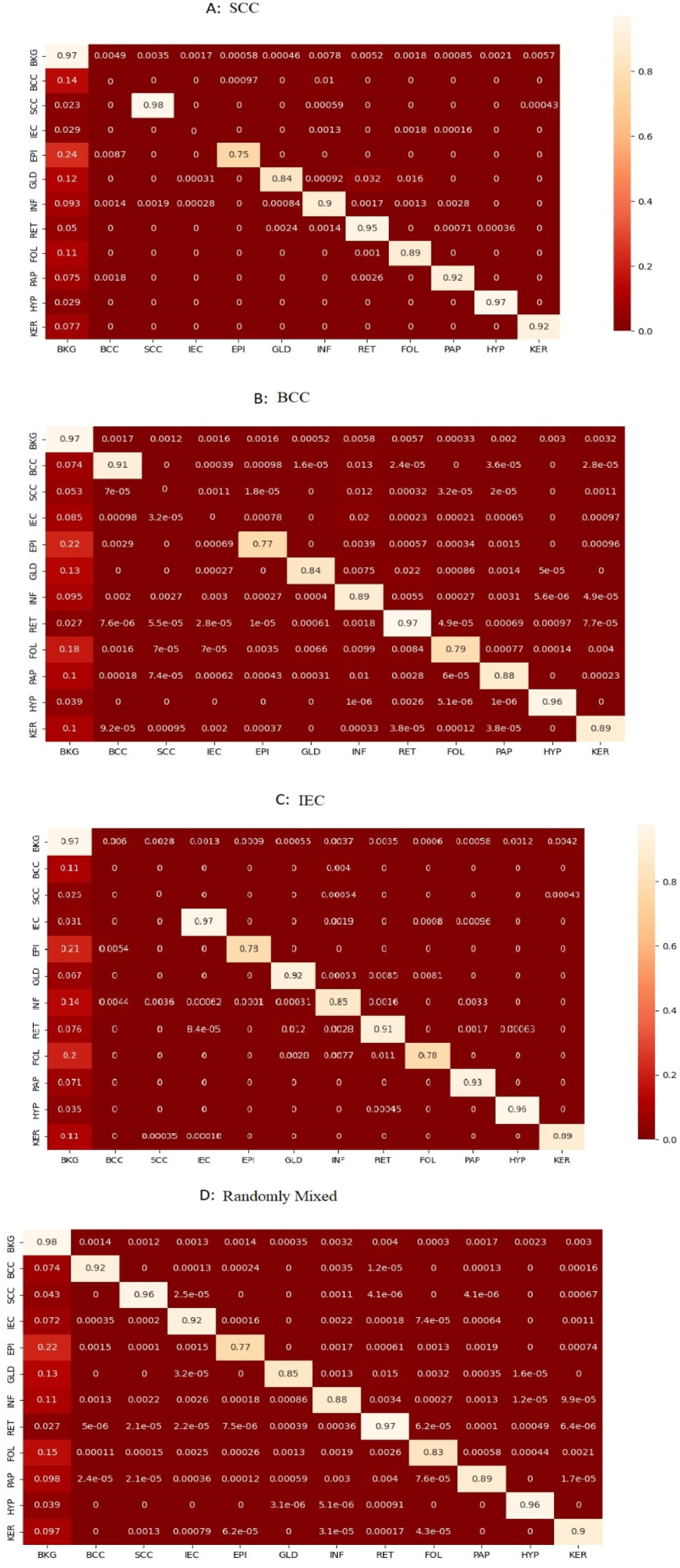
(A) The normalized confusion matrix for squamous cell carcinoma (SCC) test samples. (B) The confusion matrix for Basal cell carcinoma (BCC) test samples. (C) The normalized confusion matrix for intraepidermal carcinoma (IEC) test samples. (D) The normalized confusion matrix for all test samples.

The heatmaps indicate regions of interest related to certain classes (e.g., BCC, IEC, inflammation). The heatmap is a visual representation created by the model or process for each class to show activation intensity. Fig 9 shows the input images, their masks, and heatmaps for several categories.

**Fig. 9.**
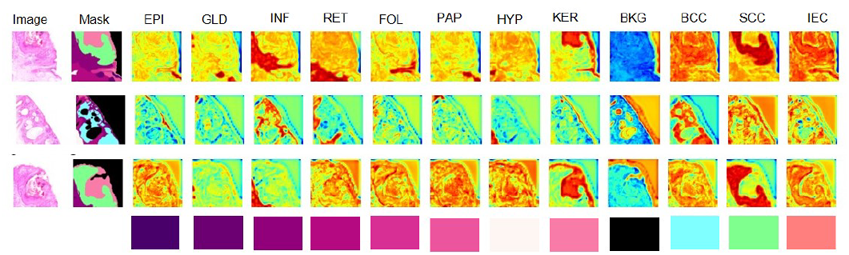
Visualize the Heatmaps produced for several classes using our incremental segmentation approach, indicating high activations, the color intensities reveal the model’s prediction confidence.

### C. Comparative Results

Our proposed framework, which incorporates incremental learning, demonstrates improved segmentation performance compared to the approach by Thomas et al. [32], which employs a UNet-based architecture for segmenting skin tissue samples into distinct layers. As shown in Table IV, our baseline Segformer model achieves higher average accuracies across different tissue categories and resolutions, indicating its superior ability to capture fine-grained histopathological structures while maintaining consistency across magnifications.

A key methodological distinction between the two approaches lies in the learning strategy. Thomas et al. use a traditional supervised learning with UNet, training three separate models where each model is capable of segmenting single single-resolution image (10X, 5X, and 2X). Instead of training distinct models, our approach gradually incorporates new resolution while keeping and refining knowledge derived from previously acquired resolutions. This allows the model to avoid catastrophic forgetting and improves generalization across several magnifications, resulting in superior segmentation accuracy on complex histological structures.

The performance gains of our baseline model over Thomas et al. [32] are shown in Table IV and are evident across all resolutions (10X, 5X, and 2X). We used the same dataset from Queensland University and performed similar preprocessing, data augmentation, and dataset splits. Data were divided into 80% training, 10% testing, and 10% validation sets to preserve class balance at all magnifications. Both our Segformer-based model and Thomas et al. technique were assessed using the same tissue classifications and resolution levels.

**TABLE IV.**
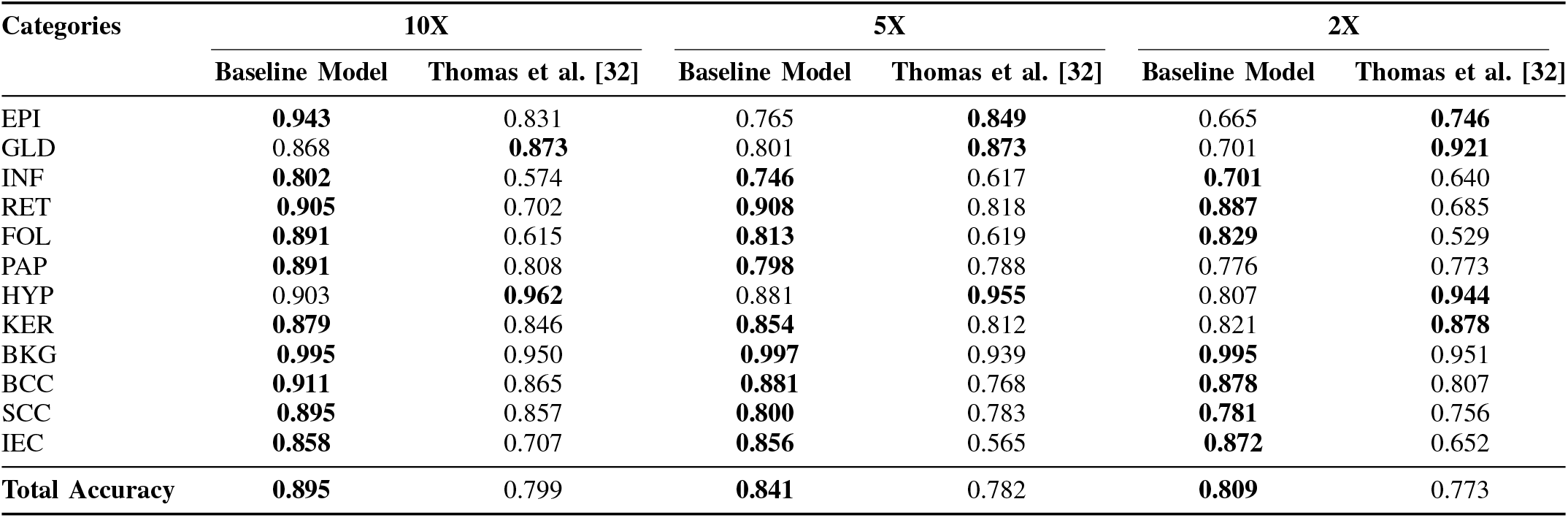
Comparison of baseline model classwise segmentation average accuracy and total accuracy with Thomas et al. [32] across the different resolutions.

For instance, at 10X resolution, our model outperforms Thomas et al. in key categories such as BCC (0.911 vs. 0.865), EPI (0.943 vs. 0.831), RET (0.905 vs. 0.702), and INF (0.802 vs. 0.574). Similar trends are observed at 5X and 2X resolutions, where our model maintains higher precision, particularly in categories like KER (0.879 vs. 0.846) and RET (0.908 vs. 0.818). These improvements highlight the effectiveness of the proposed incremental learning approach in enhancing segmentation accuracy and robustness across varying resolutions.

While our Segformer-based model generally outperforms Thomas et al., there were a few exceptions for particular tissue classes such as GLD (Granular Layer Degradation), HYP (Hyperkeratosis), and KER (Keratin), where the U-Net approach performed better at certain resolutions. This can be due to architectural differences: Segformer’s Transformer backbone excels at collecting global contextual information but can overlook fine-grained in texture-specific patterns that are critical for these classes. In contrast, U-Net’s convolutional layout is more suited to capturing local textures and intensity variations from these specific classes. Besides, there could also be cases of unequal data coverage contributing to that. For example, the classes such as HYP and GLD may not be sufficiently available in the basis dataset as a whole for the model to generalize sufficiently from those classes to others. This is seen mostly at both 5X and 2X resolution counts, where the difference between these pixels is huge.

Table V shows that our proposed model outperforms Imran et al. [33] in every category. The results of this study show that the incremental learning segmentation approach outperforms all other models in terms of segmentation accuracy. In short, our findings illustrate the incremental learning framework’s ability to achieve greater accuracy in segmentation at various resolutions, illustrating its efficacy as a reliable strategy for clinical imaging segmentation.

**TABLE V.**
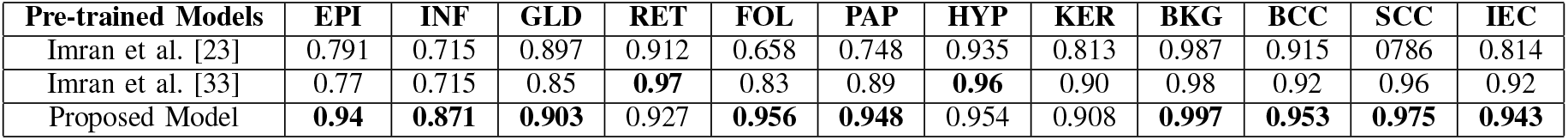
A comparison of segmentation accuracy across multiple models demonstrates the enhanced efficiency of the data incremental architecture at various resolutions.

| Pre-trained Models | EPI | INF | GLD | RET | FOL | PAP | HYP | KER | BKG | BCC | SCC | IEC |
| --- | --- | --- | --- | --- | --- | --- | --- | --- | --- | --- | --- | --- |
| Imran et al. [23] | 0.791 | 0.715 | 0.897 | 0.912 | 0.658 | 0.748 | 0.935 | 0.813 | 0.987 | 0.915 | 0.786 | 0.814 |
| Imran et al. [33] | 0.77 | 0.715 | 0.85 | **0.97** | 0.83 | 0.89 | **0.96** | 0.90 | 0.98 | 0.92 | 0.96 | 0.92 |
| Proposed Model | **0.94** | **0.871** | **0.903** | 0.927 | **0.956** | **0.948** | 0.954 | 0.908 | **0.997** | **0.953** | **0.975** | **0.943** |

The proposed incremental learning technique effectively addresses catastrophic forgetting and improves segmentation performance across resolutions, as demonstrated by the forgetting and accuracy retention curves in Figure 10. The forgetting curve, determined by IoU scores, starts at 0.802 after training on 10X images, increases to 0.853 after incorporating 5X data (Step 1), and then rises to 0.887 after integrating 2X data (Step 2). This steady enhancement demonstrates that the model successfully maintains previously learned knowledge while also retaining new information. Similarly, the accuracy improves from 0.895 in the first step to 0.926 in Step 1 and 0.955 in Step 2, indicating a significant generalization across magnifications. These enhancements imply that knowledge distillation helps the model to adjust to multiscale traits efficiently. In general, the findings support the suggested approach for whole slide image (WSI) segmentation, where learning between magnifications is critical to accurate tissue and detection of diseases.

**Fig. 10.**
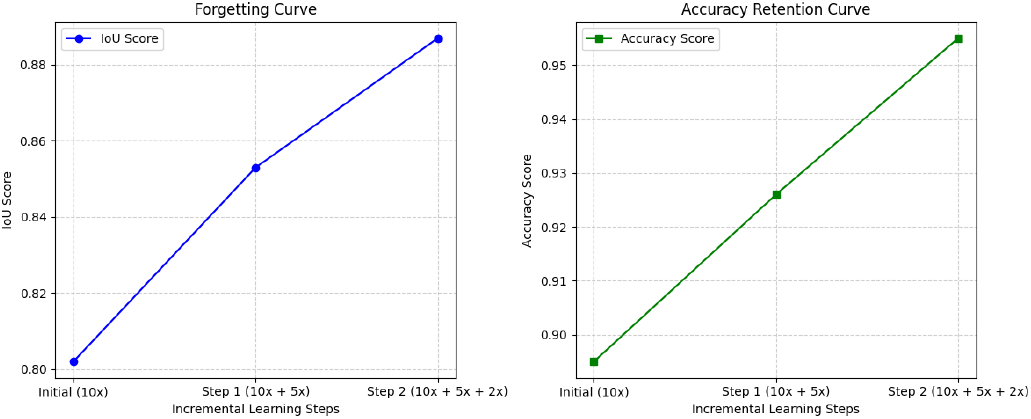
Forgetting and Accuracy Retention Curves: Evaluating the Impact of Incremental Learning on Segmentation Performance Across Multiple Magnifications

Learning different magnifications allows for precise tissue and disease characterization while maintaining previously learned information and gradually improving segmentation performance, making it ideal for multiresolution medical image analysis.

The suggested model outperforms the two earlier models by Imran et al. [33], [23] in terms of segmentation accuracy across diverse skin cancer types. Specifically, the suggested model exhibits improved accuracy in classes such as BCC, SCC, and EPI, with BCC at 0.953, SCC at 0.975, and EPI at 0.940. Notably, the model excels at identifying malignant areas, particularly in difficult categories such as HYP and PAP, where it scores 0.954 and 0.948 accuracy, accordingly, surpassing the other models by a wide margin. These results illustrate the efficiency of the suggested methodology in providing accurate and trustworthy segmentation for varied skin cancer types.

## VI. Conclusion and Future Directories

One of the biggest challenges for deep learning models is adapting and improving over time. Incremental learning solves this by enabling models to grow and adapt through successive learning stages. The proposed structure is an adaptable approach for studying histopathology images, allowing the segmentation of multiple groups within them. An incremental learning architecture has been included to improve the system’s flexibility and adaptability, with a focus on addressing the constraints presented by limited labeled data in healthcare diagnostics. Innovative transformer-based models, such as Segformer and ViT, act as the foundation for segmentation tasks, ensuring cutting-edge performance. The model provides interpretability by showing uncertainty and attention maps, which complements its segmentation skills. When compared to previous benchmark research in the field, the proposed framework outperformed them. The study focuses on the connection between model performance and data resolution, giving insights for optimizing for better results. In future, the suggested framework can be evaluated using skin cancer histology images as a major use case. It has a potential to be used for histopathology images of diverse biopsy tissues to assess the performance.

## Data Availability

The dataset analyzed in this study was provided by Queensland University and is available upon request from the data providers. All additional results generated during this study are contained within the manuscript.

## Data availability Statement

The dataset used and analyzed during the current study “Histopathology Non-Melanoma Skin Cancer Segmentation Dataset” is publicly available at https://espace.library.uq.edu.au/view/UQ:8be4bd0

## Acknowledgement

We would like to acknowledge the financial support from the MITACS Global Research Award (GRA) for making this research possible. Their funding has been instrumental in advancing this project and facilitating valuable international collaboration.

## author contributions statement

S.F. and S.B.A conceptualized and designed the study, and led the overall research process, also contributed to data interpretation and manuscript drafting with M.U.A. The data was statistically analyzed by A.A.S. The author M.U.A assisted in writing the methodology section and reviewed the manuscript.

S.F and A.A.S developed the research tools and instruments used for data augmentation and contributed to revision of the manuscript. I.A.H provided critical feedback on the research design and methodology. S.B.A contributed to writing the results section and manuscript review.

## References

[1] N. Fullman et al., “Measuring progress and projecting attainment on the basis of past trends of the health-related Sustainable Development Goals in 188 countries: an analysis from the Global Burden of Disease Study 2016,” The Lancet, vol. 390, no. 10100, pp. 1423–1459, 2017.

[2] J. M. Kocarnik *et al*., “Cancer incidence, mortality, years of life lost, years lived with disability, and disability-adjusted life years for 29 cancer groups from 2010 to 2019: a systematic analysis for the global burden of disease study 2019,” *JAMA Oncol.*, vol. 8, no. 3, pp. 420–444, 2022.

[3] P. Aggarwal, P. Knabel, and A. B. Fleischer Jr, “United States burden of melanoma and non-melanoma skin cancer from 1990 to 2019,” *J. Am. Acad. Dermatol.*, vol. 85, no. 2, pp. 388–395, 2021.

[4] R. L. Siegel, A. N. Giaquinto, and A. Jemal, “Cancer statistics, 2024,” *CA Cancer J. Clin.*, vol. 74, no. 1, pp. 12–49, 2024.

[5] M. A. Hayat, *Brain Metastases from Primary Tumors, Volume 3: Epidemiology, Biology, and Therapy of Melanoma and Other Cancers*. Academic Press, 2016.

[6] A. T. Feldman and D. Wolfe, “Tissue processing and hematoxylin and eosin staining,” *Histopathology: Methods and Protocols*, pp. 31–43, 2014, Springer.

[7] A. H. Fischer, K. A. Jacobson, J. Rose, and R. Zeller, “Hematoxylin and eosin staining of tissue and cell sections,” *Cold Spring Harbor Protocols*, vol. 2008, no. 5, pp. pdb–prot4986, 2008, Cold Spring Harbor Laboratory Press.

[8] W. Gouda, N. U. Sama, G. Al-Waakid, M. Humayun, and N. Z. Jhanjhi, “Detection of skin cancer based on skin lesion images using deep learning,” in *Healthcare*, vol. 10, no. 7, p. 1183, 2022, MDPI.

[9] R. P. Singh, G. L. Hom, M. D. Abramoff, J. P. Campbell, M. F. Chiang, et al., “Current challenges and barriers to real-world artificial intelligence adoption for the healthcare system, provider, and the patient,” Transl. Vis. Sci. Technol., vol. 9, no. 2, pp. 45–45, 2020.

[10] R. C. Maron, J. S. Utikal, A. Hekler, A. Hauschild, E. Sattler, et al., “Artificial intelligence and its effect on dermatologists’ accuracy in dermoscopic melanoma image classification: web-based survey study,” J. Med. Internet Res., vol. 22, no. 9, p. e18091, 2020.

[11] S. Wang, B. Kang, J. Ma, X. Zeng, M. Xiao, et al., “A deep learning algorithm using CT images to screen for Corona Virus Disease (COVID-19),” Eur. Radiol., vol. 31, pp. 6096–6104, 2021.

[12] P. Ström, K. Kartasalo, H. Olsson, L. Solorzano, B. Delahunt, et al., “Artificial intelligence for diagnosis and grading of prostate cancer in biopsies: a population-based, diagnostic study,” Lancet Oncol., vol. 21, no. 2, pp. 222–232, 2020.

[13] T. J. Brinker, A. Hekler, A. H. Enk, J. Klode, A. Hauschild, et al., “Deep learning outperformed 136 of 157 dermatologists in a head-to-head dermoscopic melanoma image classification task,” Eur. J. Cancer, vol. 113, pp. 47–54, 2019.

[14] J. Leo and J. Kalita, “Survey of continuous deep learning methods and techniques used for incremental learning,” Neurocomputing, vol. 582, p. 127545, 2024.

[15] Z. Li and D. Hoiem, “Learning without forgetting,” IEEE Trans. Pattern Anal. Mach. Intell., vol. 40, no. 12, pp. 2935–2947, 2017.

[16] P. Dhar, R. V. Singh, K.-C. Peng, Z. Wu, and R. Chellappa, “Learning without memorizing,” in Proc. IEEE/CVF Conf. Comput. Vis. Pattern Recognit., 2019, pp. 5138–5146.

[17] Y. Wu, Y. Chen, L. Wang, Y. Ye, Z. Liu, et al., “Incremental classifier learning with generative adversarial networks,” arXiv preprint 1802.00853, 2018.

[18] F. Cermelli, M. Mancini, S. R. Bulo, E. Ricci, and B. Caputo, “Modeling the background for incremental learning in semantic segmentation,” in Proc. IEEE/CVF Conf. Comput. Vis. Pattern Recognit., 2020, pp. 9233– 9242.

[19] U. Michieli and P. Zanuttigh, “Incremental learning techniques for semantic segmentation,” in Proc. IEEE/CVF Int. Conf. Comput. Vis. Workshops, 2019, pp. 0–0.

[20] F. Cermelli, D. Fontanel, A. Tavera, M. Ciccone, and B. Caputo, “Incremental learning in semantic segmentation from image labels,” in Proc. IEEE/CVF Conf. Comput. Vis. Pattern Recognit., 2022, pp. 4371– 4381.

[21] E. Xie, W. Wang, Z. Yu, A. Anandkumar, J. M. Alvarez, and P. Luo, “SegFormer: Simple and efficient design for semantic segmentation with transformers,” Adv. Neural Inf. Process. Syst., vol. 34, pp. 12077–12090, 2021.

[22] G. M. Kosgiker and A. Deshpande, “A novel SEGCAP algorithm based enhanced segmentation of dermoscopic images of interest,” Mater. Today Proc., vol. 51, pp. 779–787, 2022.

[23] M. Imran, M. I. Tiwana, M. M. Mohsan, N. S. Alghamdi, and M. U. Akram, “Transformer-based framework for multi-class segmentation of skin cancer from histopathology images,” Front. Med., vol. 11, p. 1380405, 2024.

[24] N. Hameed, A. M. Shabut, M. K. Ghosh, and M. A. Hossain, “Multiclass multi-level classification algorithm for skin lesions classification using machine learning techniques,” Expert Syst. Appl., vol. 141, p. 112961, 2020.

[25] N. Moradi and N. Mahdavi-Amiri, “Multi-class segmentation of skin lesions via joint dictionary learning,” Biomed. Signal Process. Control, vol. 68, p. 102787, 2021.

[26] A. Vaswani, “Attention is all you need,” in Adv. Neural Inf. Process. Syst., 2017.

[27] A. Dosovitskiy et al., “An image is worth 16×16 words: Transformers for image recognition at scale,” arXiv preprint 2010.11929, 2020.

[28] W. Wang, E. Xie, X. Li, D.-P. Fan, K. Song, et al., “Pyramid vision transformer: A versatile backbone for dense prediction without convolutions,” in Proc. IEEE/CVF Int. Conf. Comput. Vis., 2021, pp. 568–578.

[29] M. A. Islam, S. Jia, and N. D. B. Bruce, “How much position information do convolutional neural networks encode?,” arXiv preprint 2001.08248, 2020.

[30] X. Chu, Z. Tian, B. Zhang, X. Wang, and C. Shen, “Conditional positional encodings for vision transformers,” arXiv preprint 2102.10882, 2021.

[31] Y. Song, P. Zhang, W. Huang, Y. Zha, T. You, and Y. Zhang, “‘Parallelcircuitized’ distillation for dense object detection,” Displays, p. 102587, 2023.

[32] S. M. Thomas, J. G. Lefevre, G. Baxter, and N. A. Hamilton, “Interpretable deep learning systems for multi-class segmentation and classification of non-melanoma skin cancer,” Med. Image Anal., vol. 68, p. 101915, 2021.

[33] M. Imran, M. U. Akram, M. I. Tiwana, A. A. Salam, T. Hassan, and D. Greco, “Two-dimensional hybrid incremental learning (2DHIL) framework for semantic segmentation of skin tissues,” Image Vis. Comput., vol. 148, p. 105147, 2024.

